# Knowledge, Attitudes, and Practices Regarding Maternal Nutrition Counselling Among Frontline Health Workers in Udupi, Karnataka, India: A Sequential Explanatory Mixed-Methods Study

**DOI:** 10.64898/2026.06.17.26355890

**Authors:** M S Amrutha, Chythra R Rao, Suvarna Hebbar, J Vennila, G. Arun Maiya, Prashanth Bhat, Namratha Pai Kotebagilu, Ramya Bhagavath, Ekta Rupani, Trisha Maji, Shashikala K Bhat

## Abstract

**Background:** India’s maternal nutrition profile is undergoing a dual-direction shift, with persistent undernutrition coexisting alongside rising overweight and micronutrient deficiencies. Despite national efforts through Integrated Child Development Services (ICDS) and the National Health Mission (NHM), maternal dietary diversity remains suboptimal in India. Frontline health workers (FLWs) play a central role in delivering nutrition counselling; however, gaps remain between knowledge and its translation into practice, highlighting the need to strengthen training, applied competencies, and health system support within primary care settings.

**Objective:** To assess knowledge, attitudes, and practices (KAP) regarding maternal nutrition counselling among FLWs and to explore contextual factors influencing counselling delivery.

**Methods:** A sequential explanatory mixed-methods study was conducted in Udupi, Karnataka, India. In phase one, 46 FLWs- Accredited Social Health Activists (ASHA), Community Health Officers (CHO), and Primary Health Care Officers (PHCO) completed a validated Knowledge, Attitudes, and Practices (KAP) questionnaire. Data were analysed using descriptive statistics, Kruskal–Wallis test, Spearman correlation, and exploratory multiple linear regression. In phase two, one focus group discussion with 21 participants was conducted and analysed using reflexive thematic analysis.

**Results:** FLWs demonstrated moderate KAP scores (37.50 ± 5.09), with lower scores observed in dietary diversity knowledge and counselling practices. CHOs and PHCOs had significantly higher knowledge (p < 0.001) and practice scores (p = 0.002) compared to ASHAs, while attitudes were similar across cadres. Knowledge was positively associated with practice (ρ = 0.389, p = 0.008). Exploratory regression indicated that cadre and knowledge were associated with practice, while attitude was not statistically significant. Qualitative findings suggested that counselling was largely protocol-based and constrained by workload, limited counselling tools, economic barriers, and cultural food practices.

**Conclusion:** Despite positive attitudes towards maternal nutrition counselling, frontline health workers demonstrated gaps in knowledge and counselling practices. Mixed-methods findings suggest that counselling delivery is shaped by both provider competencies and health-system constraints, highlighting the need for implementation-focused strategies to strengthen maternal nutrition counselling in routine antenatal care.

## Introduction

Maternal nutrition in India is undergoing a profound epidemiological transition, shifting from focus on caloric and macronutrient deficits to the complex phenomenon known as the Triple Burden of Malnutrition(1). This contemporary paradigm is characterised by the simultaneous, often overlapping, coexistence of chronic undernutrition, hidden hunger (micronutrient deficiencies), and a rapidly rising prevalence of overnutrition and obesity(2)(3). This transition reflects a shifting disease burden in women of reproductive age from undernutrition to a complex coexistence of multiple nutritional risks.

The nutritional status of expectant mothers continues to be a critical determinant of intergenerational health and societal development(4).India has made significant progress in reducing maternal mortality, with the rate declining from 481 per 100,000 births in 1990 to 99 in 2020(5)(6). However, the nutritional status of pregnant women remains critical, as only 18% meet dietary diversity recommendations. On average, Indian women gain 7 kg of weight during pregnancy, against the recommendation of 10-12 kg(7). The prevalence of maternal low BMI (<18.5 kg/m^2^) is 10%-20% globally, but 30%-40% in South Asia and India (8). Currently, 57% of women of reproductive age are anaemic, and 19% are underweight, with only 44 % of women consuming IFA for the recommended 100 days(9)(10). Recent national data indicate that nearly 24% of women of reproductive age in India are overweight or obese, with higher prevalence in states such as Punjab and southern regions including Tamil Nadu, Karnataka, and Kerala, highlighting clear regional clustering and a growing public health concern. Collectively, these indicators reflect a persistent coexistence of undernutrition, micronutrient deficiencies, and emerging overnutrition among Indian women, indicating a complex nutritional transition requiring context-specific and systems-level interventions(11).

In many low- and middle-income countries (LMICs), pregnant women’s diets are often inadequate, with limited consumption of micronutrient-dense foods such as non-vegetarian items, fruits, and vegetables(12). Socio-economic, cultural, and systemic factors further complicate maternal nutrition. Factors like education, household decision-making power, access to resources, and traditional beliefs influence dietary behaviours (13). Traditional beliefs and misconceptions also act as a hindrance, as women avoid certain nutritious foods during pregnancy, as they believe it could lead to congenital disorders in newborns(14).

In response to these challenges, several government initiatives and health programmes like the Integrated Child Development Services (ICDS) and the National Health Mission (NHM) have been brought into action to improve maternal and child health. These programs depend on frontline health workers to provide services such as supplementary nutrition, antenatal care (ANC), and nutrition counselling (15). All women, regardless of nutritional status, should be provided with assistance on 1. Screening for Nutritional Risk. 2. Iron and Folic Acid Supplementation. 3. Healthy eating practices such as consumption of iodised salt, reducing caffeine intake, restricting tobacco, alcohol and other toxic substances. 4. Deworming <u>[24]</u>.

Frontline workers (FLWs) serve as a vital link between the healthcare system and remote communities, ensuring access to essential services(16). Their close relationship with these populations allows them to understand local dietary habits and cultural practices, which helps improve maternal nutrition outcomes(17). Maternal malnutrition deaths could decrease by 15% if nutrition-focused interventions reach 90% of the population. Effective delivery of these interventions to those in need of care could prevent approximately 28% of maternal and neonatal deaths and 22% of stillbirths(18)(19).

Despite numerous efforts, shortcomings remain in the proper implementation of these programmes. The execution of maternal nutrition counselling and dietary diversity remains inconsistent, often failing to bridge the gap between biomedical recommendations and community practices(20). The performance of FLWs is often hindered by a lack of supervision, resources, infrastructure, fair honoraria, and adequate training and helpers, leading to a heavier workload(21). Implementation challenges arise when there is a disconnect between training and real-world application. Counselling is often not tailored to the specific needs of women and fails to adapt to their social and cultural backgrounds, which reduces its effectiveness (22). Successful implementation would require adapting to local circumstances, constant monitoring and managing systemic barriers as emphasised by the Knowledge-to-Action framework(23). Discrepancies between recommendations and personal practices can undermine credibility and effectiveness(24).

Nutrition counselling has emerged as a key strategy to improve dietary behaviours among pregnant and lactating women. Behaviour change communication interventions have shown promising results in enhancing dietary diversity and improving maternal and child health outcomes(25).

This study provides an integrated assessment of frontline health workers’ knowledge, attitudes, and practices regarding maternal nutrition counselling. Specifically, it aims to assess knowledge, attitudes, and practices related to maternal nutrition counselling and to explore counselling practices, implementation barriers, and facilitators influencing dietary diversity using a sequential explanatory mixed-methods design.

## Methodology

This study adopted a sequential explanatory mixed-methods design, with an initial quantitative phase followed by a qualitative phase. The quantitative component evaluated the knowledge, attitudes, and practices related to maternal nutrition counselling and dietary diversity among frontline health workers, while the qualitative component was conducted to contextualise and deepen understanding of the quantitative results. The study was carried out in selected Primary Health Centres and Urban Primary Health Centres in Udupi taluk, Karnataka, India.

Participants included frontline health workers involved in antenatal care services, namely Community Health Officers (CHO), Primary Health Care Officers (PHCO), and Accredited Social Health Activists (ASHA) working in the study area. Ethical approval was obtained from the Institutional Ethics Committee (IEC No. IEC1:379/2024), and the study was registered with the Clinical Trials Registry of India (CTRI No. CTRI/2024/11/076863).Administrative approval was obtained from the Department of Health and Family Welfare, Udupi District prior to data collection.). Participant recruitment and data collection for the quantitative phase were conducted between 25 March 2025 and 23 June 2025. The qualitative phase, comprising one focus group discussion, was conducted on 23 October 2025.This investigation formed the baseline formative phase of a larger capacity-building intervention aimed at informing the development of strategies to strengthen maternal nutrition counselling practices among frontline health workers. The quantitative component was reported in accordance with the STROBE(S4 File) guidelines for cross-sectional studies, and the qualitative component was reported in accordance with the COREQ(S5 File) guidelines for qualitative research. Methodological rigor of the mixed-methods design was additionally appraised using the Mixed Methods Appraisal Tool (MMAT, 2018). Integration of quantitative and qualitative findings was achieved through a joint display approach, with results synthesized in Table 5 to identify convergence between datasets during the interpretation phase. The qualitative phase was designed to explain and contextualize quantitative findings demonstrating variation in knowledge and counselling practices among cadres.

### Phase 1: Quantitative Study

#### Study Design

A cross-sectional study design was adopted for the quantitative phase. The total number of eligible healthcare providers was a defined population consisting of 32 Community Health Officers, 43 Primary Health Care Officers, and 157 Accredited Social Health Activists. As this study was intended as an exploratory formative assessment to inform the development of a subsequent capacity-building intervention, a pragmatic sample was selected to ensure representation across frontline health worker cadres while maintaining operational feasibility. The study was designed to generate preliminary evidence regarding maternal nutrition counselling practices rather than to test predefined hypotheses. Participants were recruited during routine monthly meetings across selected health centres where all eligible frontline health workers were present. The final sample comprised 6 Community Health Officers, 9 Primary Health Care Officer, and 31 Accredited Social Health Activists. Participants were selected from multiple centres, including Kemmannu, Kukkehalli, Manipal, Pernankila, and Udupi, ensuring representation from both rural and urban settings. Within the selected centres, participants were recruited during the routine monthly meetings, which allowed access to all eligible healthcare providers present at the time.

#### Inclusion Criteria

Frontline healthcare workers currently providing services in Udupi taluk and those willing to participate and provide informed consent were included in the study.

#### Data Collection

Data were collected using a semi-structured Knowledge, Attitude, and Practice(KAP) questionnaire focusing on maternal nutrition and dietary diversity. The KAP questionnaire (S1 File) was developed based on study objectives, relevant literature, and national guidelines. Items covered dietary diversity, balanced diets, micronutrient nutrition, nutritional risk assessment, BMI classification, gestational weight gain recommendations, anaemia prevention, supplementation practices, and dietary management of common pregnancy-related conditions.Content validity was established through expert review by a multidisciplinary panel with expertise in maternal nutrition, public health, community medicine, and obstetrics. The Content Validity Index (CVI) demonstrated good agreement among experts, with an overall Scale-level CVI (S-CVI) of 0.89. Internal consistency reliability was acceptable, with a Cronbach’s alpha of 0.709.The questionnaire included sociodemographic details and 48 items assessing knowledge, attitude, and practices (KAP) related to maternal nutrition counselling. Knowledge and practice items were scored as 1 for correct and 0 for incorrect responses, while attitude items were scored on a 3-point Likert scale (1–3), with higher scores indicating a more positive attitude. The questionnaire was administered in the local language, and participants were given approximately 45 minutes to complete it. To minimise information bias, participants completed questionnaires independently and were instructed not to discuss responses with colleagues during data collection. Recruitment was conducted across multiple centres to improve representation across cadres.

#### Data Analysis

All questionnaires were reviewed for completeness at the time of collection. No missing data were identified; therefore, complete-case analysis was performed. Statistical analysis was performed using descriptive and inferential methods. Descriptive statistics, including mean, standard deviation, and 95% confidence intervals, were used to summarise socio-demographic characteristics and knowledge, attitude, and practice (KAP) scores. As the data were not normally distributed, the Kruskal–Wallis test was used to compare KAP scores across professional groups (ASHA, CHO, and PHCO). Where significant differences were identified, Dwass–Steel–Critchlow–Fligner (DSCF) pairwise comparisons were performed to identify group-wise differences. Effect size was calculated using epsilon squared (ε²). Correlation between KAP domains was assessed using Spearman’s rank correlation coefficient. For regression analysis, categorical variables were dummy coded with ASHA as the reference category. Multiple linear regression was performed to identify potential predictors of KAP scores. A p-value of <0.05 was considered statistically significant.

#### Result

**Table 1.** : Socio-demographic characteristics of participants (n = 46)

| Variable |  | ASHA<br>(n=31) | CHO (n=6) | PHCO<br>(n=9) | Total |
| --- | --- | --- | --- | --- | --- |
| Age (years) | | 45.87 ± 7.79 | 31.83 ± 5.23 | 33.33 ± 5.85 | 41.59 ± 9.41 |
| Work experience (years) | | 7.82 ± 4.28 | 4.28 ± 1.23 | 6.33 ± 3.74 | 7.07 ± 4.05 |
| Education | Secondary stage | 27 (87.1) | 0 (0.0) | 7 (77.8) | 34 (73.9) |
|  | Undergraduate | 1 (3.2) | 5 (83.3) | 1 (11.1) | 7 (15.2) |
|  | Postgraduate | 0 (0.0) | 1 (16.7) | 1 (11.1) | 2 (4.3) |
|  | Middle stage | 2 (6.5) | 0 (0.0) | 0 (0.0) | 2 (4.3) |
|  | Preparatory stage | 1 (3.2) | 0 (0.0) | 0 (0.0) | 1 (2.2) |

**Note: Educational qualifications were categorized as foundational (pre-school to Class 2), preparatory (Classes 3–5), middle (Classes 6–8), secondary (Classes 9–12), undergraduate, postgraduate, and doctoral levels.**

ASHA workers were older and had higher work experience compared to CHOs and PHCOs. Educational attainment varied across cadres, with CHOs having higher levels of undergraduate and postgraduate education compared to ASHAs and PHCOs.

The overall KAP score was 37.50 ± 5.09 out of the maximum attainable score, indicating moderate levels of knowledge, attitude, and practice among frontline health workers.However, domain-wise analysis revealed an uneven distribution across knowledge, attitude, and practice components rather than uniform performance.

**Table 2.** Domain-wise Assessment of Knowledge, Attitude, and Practice in Maternal Nutrition Counselling among Frontline Health Workers.

| Domain | Sub-domain | Item description | Mean percentage of correct responses (%) |
| --- | --- | --- | --- |
| **Knowledge** | Balanced diet understanding | Components of a balanced diet during pregnancy | 17.4 |
|  | Dietary diversity | Minimum dietary diversity recommendations (MDD-W concept) | 19.6 |
|  | Macronutrient knowledge | Identification of food groups and protein sources | 21.8 |
|  | Micronutrient knowledge | Iron, folate, iodine-rich foods and anaemia prevention | 23.9 |
|  | Nutritional status assessment | BMI classification and gestational weight gain recommendations | 24.7 |
|  | Pregnancy-related dietary conditions | Nutrition in constipation, nausea, and related conditions | 27.3 |
| **Practice** | Counselling frequency | Routine antenatal nutrition counselling delivery | 23.9 |
|  | Dietary counselling content | Advice on balanced diet and dietary diversity | 25.1 |
|  | Symptom management counselling | Nausea, bloating, constipation dietary advice | 27.8 |
|  | Supplement counselling | Iron–folic acid timing, absorption, and adherence support | 24.6 |
|  | Behaviour change counselling | Reinforcement of healthy dietary practices | 26.2 |
| **Attitude** | Perceived importance | Value of nutrition counselling in antenatal care | 68.5 |

**Note:** Percentages represent the mean proportion of correct responses across questionnaire items within each sub-domain. Individual questionnaire items are provided in S1 File.

Knowledge scores demonstrated marked deficiencies in foundational maternal nutrition concepts, with low correct response rates in balanced diet understanding (17.4%) and dietary diversity (19.6%). Similar low performance was observed in micronutrient-related knowledge (23.9%) and nutritional status assessment (24.7%), indicating consistent gaps in essential maternal nutrition knowledge.

Practice-related findings demonstrated gaps in counselling delivery, particularly in routine counselling frequency (23.9%) and iron–folic acid counselling practices (24.6%), suggesting limited translation of knowledge into practice. Behaviour change counselling and dietary counselling practices also remained limited (25–26%), indicating suboptimal implementation of maternal nutrition counselling within routine antenatal care.

Conversely, attitude scores were relatively high (68.5%), indicating that frontline health workers recognised and valued the importance of maternal nutrition counselling despite gaps in knowledge and practice. These findings suggest a potential knowledge–practice gap, whereby positive attitudes were not consistently reflected in counselling practices.

Comparison of knowledge, attitude, and practice (KAP) scores across professional cadres of frontline health workers is presented in **Table 3**. Knowledge scores showed a statistically significant difference between groups (p < 0.001), with CHOs and PHCOs scoring higher than ASHA workers. Practice scores also differed significantly across groups (p = 0.002), while no significant difference was observed for attitude scores (p = 0.403).

**Table 3.** Comparison of Knowledge, Attitude, and Practice Scores across Professional Cadres of Frontline Health Workers.

| Variable | ASHA | CHO | PHCO | $\chi^2$ (Kruskal–Wallis) | df | p-value | $\epsilon^2$ |
| --- | --- | --- | --- | --- | --- | --- | --- |
| Knowledge | 14.65 ±<br>2.77 | 18.33 ±<br>3.07 | 18.11 ±<br>1.62 | 14.38 | 2 | <0.001* | 0.320 |
| Attitude | 12.29 ±<br>1.79 | 12.33 ±<br>1.96 | 13.22 ±<br>1.20 | 1.82 | 2 | 0.403 | 0.040 |
| Practice | 8.55 ±<br>2.31 | 9.83 ±<br>2.93 | 11.11 ±<br>1.17 | 13.00 | 2 | 0.002* | 0.289 |

Note: Effect size was estimated using epsilon squared ( $\epsilon^2$ ). Pairwise comparisons following significant Kruskal–Wallis tests were conducted using the Dwass–Steel–Critchlow–Fligner (DSCF) procedure.

Post-hoc pairwise comparisons indicated that ASHA workers had significantly lower knowledge scores compared to CHOs (p = 0.039) and PHCOs (p = 0.003). Similarly, ASHA workers had significantly lower practice scores compared to PHCOs (p = 0.002).

Spearman’s rank correlation analysis demonstrated a moderate positive association between knowledge and practice (ρ = 0.389, p = 0.008). No significant correlations were observed between knowledge and attitude (ρ = 0.026, p = 0.862) or between attitude and practice (ρ = 0.183, p = 0.223).

**Table 4.** Exploratory multiple linear regression analysis of factors associated with knowledge and practice scores among frontline health workers.

| Variable | Knowledge B (SE) | p-value | 95% CI | Practice B (SE) | p-value | 95% CI |
| --- | --- | --- | --- | --- | --- | --- |
| Age | 0.031 (0.058) | 0.596 | -0.084 to 0.146 | -0.022 (0.051) | 0.667 | -0.122 to 0.078 |
| Work Experience | 0.082 (0.104) | 0.433 | -0.123 to 0.287 | -0.041 (0.091) | 0.656 | -0.220 to 0.138 |
| Cadre (CHO vs ASHA) | 5.984 (1.921) | 0.003* | 2.214 to 9.754 | -1.562 (1.804) | 0.389 | -5.101 to 1.977 |
| Cadre (PHCO vs ASHA) | 3.112 (1.302) | 0.019* | 0.551 to 5.673 | 0.742 (1.210) | 0.540 | -1.630 to 3.114 |
| Total Knowledge | — | — | — | 0.287 (0.132) | 0.034* | 0.028 to 0.546 |

Note: Regression analyses were conducted for exploratory purposes. The knowledge model demonstrated moderate explanatory power ( $R^2 = 0.487$ ; adjusted $R^2 = 0.341$ ), while the practice model demonstrated limited explanatory power ( $R^2 = 0.305$ ; adjusted $R^2 = 0.106$ ).

Multiple linear regression analyses were conducted to identify factors associated with knowledge and practice scores among frontline health workers. The models were simplified to ensure statistical stability given the limited sample size.

In the knowledge model, age, work experience, and professional cadre were included as predictors. The analysis showed that the professional cadre was significantly associated with knowledge scores, with CHOs (B = 5.984, p = 0.003) and PHCOs (B = 3.112, p = 0.019) demonstrating higher knowledge scores than ASHA workers. Age and work experience were not statistically significant predictors.

In the practice model, age, work experience, professional cadre, and total knowledge score were included as variables. Total knowledge score showed a statistically significant positive association with practice scores (B = 0.287, p = 0.034), indicating that higher knowledge scores were associated with higher practice scores. Other variables were not statistically significant.

### Phase 2: Qualitative Study (Focus Group Discussion)

#### Study Design

In the second phase, a qualitative descriptive design using a Focus Group Discussion (FGD) was employed to gain in-depth insights into maternal nutrition counselling practices. Purposive sampling was used to select participants with relevant experience in antenatal care. Healthcare providers with a minimum of five years of experience were included. However, for Community Health Officers (CHOs), a minimum of one year of experience was considered sufficient due to the recent introduction of this cadre in 2018. A total of 24 participants were initially planned, in line with standard methodological recommendations for focus group discussions and to ensure representation from all three cadres. However, the final FGD included 21 participants, as three were unavailable on the day of data collection. The group comprised 8 Accredited Social Health Activists (ASHAs), 6 Primary Health Care Officers (PHCOs), and 7 Community Health Officers (CHOs). The discussion was conducted at the Primary Health Centre, Kemmannu, which was the only centre where all three cadres were sufficiently represented, allowing for a comprehensive discussion and diverse perspectives.

#### Development of FGD Guide

A semi-structured FGD guide was developed based on study objectives, findings from the quantitative phase, national maternal nutrition guidelines, dietary diversity frameworks, and relevant literature(S2 File).

The guide was reviewed by subject experts to ensure content validity and contextual relevance. It included open-ended questions covering areas such as maternal nutrition knowledge, screening practices, barriers to dietary diversity practices, counselling challenges, supplement use, resource gaps, training needs, and successful field-level practices.

#### Data Collection

The FGD was conducted in the local language in a private setting at the Primary Health Centre to ensure confidentiality and encourage open discussion. The session lasted approximately 45 minutes and was audio-visually recorded. The focus group discussion was facilitated by the female primary researcher (AMS), with prior training and experience in qualitative research. No prior research relationship was established between the facilitator and participants before data collection. Participants were informed about the objectives of the study and the role of the researcher before commencement of the discussion. The facilitator’s professional background as a clinical nutrition researcher was disclosed, and reflexive discussions were conducted within the research team throughout the process to enhance analytical rigor. Audio-recording of the focus group discussion and field-note documentation were carried out by a trained co-researcher (RB). To ensure that findings accurately reflected participants’ perspectives, key points were summarised during the discussion and participant confirmation was sought through member checking.

#### Data Analysis

We began by transcribing the audio recordings from the focus group discussion verbatim, followed by translation into English. Each translation was carefully compared with the original recordings to ensure accuracy. All personal identifiers were removed, and participants were assigned unique anonymized codes to maintain confidentiality. The finalized transcripts were imported into ATLAS.ti for systematic organization and management of the data.

Reflexive thematic analysis was conducted following the framework of Virginia Braun and Victoria Clarke, involving familiarization with the transcripts, identification of meaningful text segments, and generation of initial codes directly from the data. Coding was primarily conducted by AMS and reviewed through discussion with an experienced qualitative researcher to enhance credibility and consistency of interpretation. Related codes were then grouped into categories, from which overarching themes and sub-themes were developed(S3 File)..Coding and analysis continued until no new codes emerged from the single focus group discussion; however, findings are interpreted as exploratory and context-specific and do not claim theoretical saturation. The inclusion of multiple cadres within the same discussion facilitated cross-professional interaction, which enriched the dataset despite the single-FGD design.

#### Theme 1: Maternal Nutrition Knowledge and Counselling Practices

Nutrition counselling provided by frontline health workers focused on dietary recommendations, micronutrient awareness, supplement protocols, and disease-specific dietary advice, with emphasis on commonly consumed foods such as green leafy vegetables, cereals, eggs, pulses, and millets. Counselling largely reflected standardized antenatal nutrition messages and routine supplementation guidance, primarily based on general dietary advice and commonly promoted food groups rather than individualized dietary assessment or tailoring to specific maternal nutritional needs.Participants consistently highlighted iron-rich and balanced dietary advice as central to pregnancy care, primarily aimed at preventing anemia and supporting fetal growth:

“If they eat green leafy vegetables, anemia will not occur… the baby inside the womb will gain more weight and grow healthily.”

Participants also reported providing dietary guidance for specific conditions such as gestational diabetes and hypertension, including reducing sugar intake, limiting salt, and adjusting meal patterns according to clinical condition.Supplementation counselling was described as a structured component of antenatal care, with trimester-based instructions:

“For the first three months, folic acid. After three months, iron and folic acid… Calcium helps prevent hypertension.”

In addition, participants mentioned advising calcium intake and explaining timing considerations between iron tablets and milk or calcium consumption to improve absorption.

#### Theme 2: Maternal Nutrition Monitoring and Assessment Practices

Participants reported assessing maternal nutritional status and dietary adherence using routine clinical observations, simple anthropometric measurements, dietary inquiry, and available laboratory information. Monitoring mainly depended on easily available indicators in field settings, with weight monitoring being the most commonly used measure. However, assessment was mostly based on observation and mothers’ reported diet, with limited use of structured tools for detailed dietary evaluation

“We mainly check monthly weight gain… We check eyes and ask about symptoms.”

In addition to weight, clinical observations such as pallor, dizziness, edema, and swelling were commonly used to identify possible nutritional or health concerns during pregnancy. Where available, hemoglobin reports and scan findings were used to support assessment and follow-up.Dietary intake was primarily assessed through informal recall methods.

“We ask what they ate during the week”

Overall, nutritional assessment practices combined multiple routine sources of information, including physical observation, reported dietary intake, and available clinical reports.

#### Theme 3: Structural and Contextual Barriers to Optimal Nutrition

Structural and contextual factors were identified as key barriers affecting maternal dietary practices and the effectiveness of nutrition counselling. Service delivery related barriers such as heavy workload, multiple program responsibilities and limited time per client were highlighted as major challenges that reduced the depth and quality of dietary counselling.

Additionally, economic constraints were frequently reported, limiting access to a diverse and nutrient-rich diet, particularly the affordability of fruits and green leafy vegetables. In such situations, participants often adapted counselling by recommending locally available and affordable alternatives.

“Some cannot afford market greens… so we advise local greens like drumstick leaves.”

Physiological factors also influenced dietary intake during pregnancy. Common issues such as nausea, vomiting, reduced appetite, and side effects of iron supplementation (particularly constipation) were reported to interfere with food consumption. In addition, food intolerances and gastric discomfort further restricted dietary variety.

Cultural beliefs and family influences were noted to affect food choices, although participants observed a gradual decline in restrictive practices. Misconceptions around certain foods, were still present .

“Some hot-cold foods belives like fruits, green leafy vegetables should not be eaten… These beliefs were often reinforced by mothers-in-law and elderly family members.

Overall, food preferences, resistance to vegetable intake, and adherence to traditional dietary practices collectively influenced dietary adherence, necessitating continuous negotiation between recommended nutritional guidelines and culturally acceptable food choices.

#### Theme 4: Family and Community Engagement in Maternal Nutrition Care

Maternal nutrition counselling, as described by frontline health workers, is primarily directed towards the pregnant woman. However, participants highlighted that dietary decisions during pregnancy are strongly shaped by family members and household practices. This creates a gap between counselling provided at the facility level and the realities of the home environment, often limiting adherence to dietary recommendations.

To bridge this gap, frontline health workers reported involving family members in counselling sessions whenever possible, recognising their significant role in influencing food choices.

“If she stays at her mother’s house, we counsel there… we involve family members.”

“If at in-laws’ house, we counsel them.”

They also reported serving as a link between patients, families, and clinicians, helping to translate dietary advice and clarify misconceptions related to nutrition and pregnancy.

“We act as mediators between them and doctor.”

#### Theme 5: Behavioural and Outcome Evaluation of Counselling

Evaluation of dietary diversity counselling was largely embedded within routine care processes rather than conducted as a formal assessment. Frontline health workers described that changes in maternal dietary behaviour were mainly inferred through follow-up interactions, where shifts in food acceptance, incorporation of recommended food groups, and reduced dietary restrictions were gradually noted over time. Counselling outcomes were reflected through indirect indicators of dietary improvement, such as better perceived maternal nutrition status and positive pregnancy outcomes, including adequate maternal weight gain and healthy neonatal birth weight. These were commonly used as practical signs that dietary advice had been followed and accepted. .

“We observe their communication style and changes in them.”

“When healthy babies with good birth weight are born, mothers say following food advice helped.”

Some successful counselling outcomes were linked to timely identification of problems and tailored dietary modifications based on individual responses and intolerance patterns. “We identified dietary triggers… later she revealed Crohn’s disease.”

#### Theme 6: Capacity Building and Training Needs in Maternal Nutrition Care

Gaps in knowledge, skills, and supportive resources among frontline health workers were identified as limiting the effectiveness of maternal nutrition counselling. Participants reported inadequate preparedness in managing complex and disease-specific nutritional conditions, particularly diabetes and gestational diabetes, along with limited understanding of micronutrient metabolism and supplementation practices, as reflected in the statement below.

“We need more knowledge about diabetes, insulin, gestational diabetes…”

They also emphasized the lack of practical, user-friendly counselling tools, highlighting the need for visual aids, videos, clip charts, and OPD-based materials to improve communication and understanding, especially in high-risk cases .

“Five-minute videos are useful, especially for high-risk cases”

Overall, findings indicate the need for continuous capacity building supported by appropriate educational resources to strengthen counselling effectiveness in routine maternal nutrition care.

**Fig 1.**
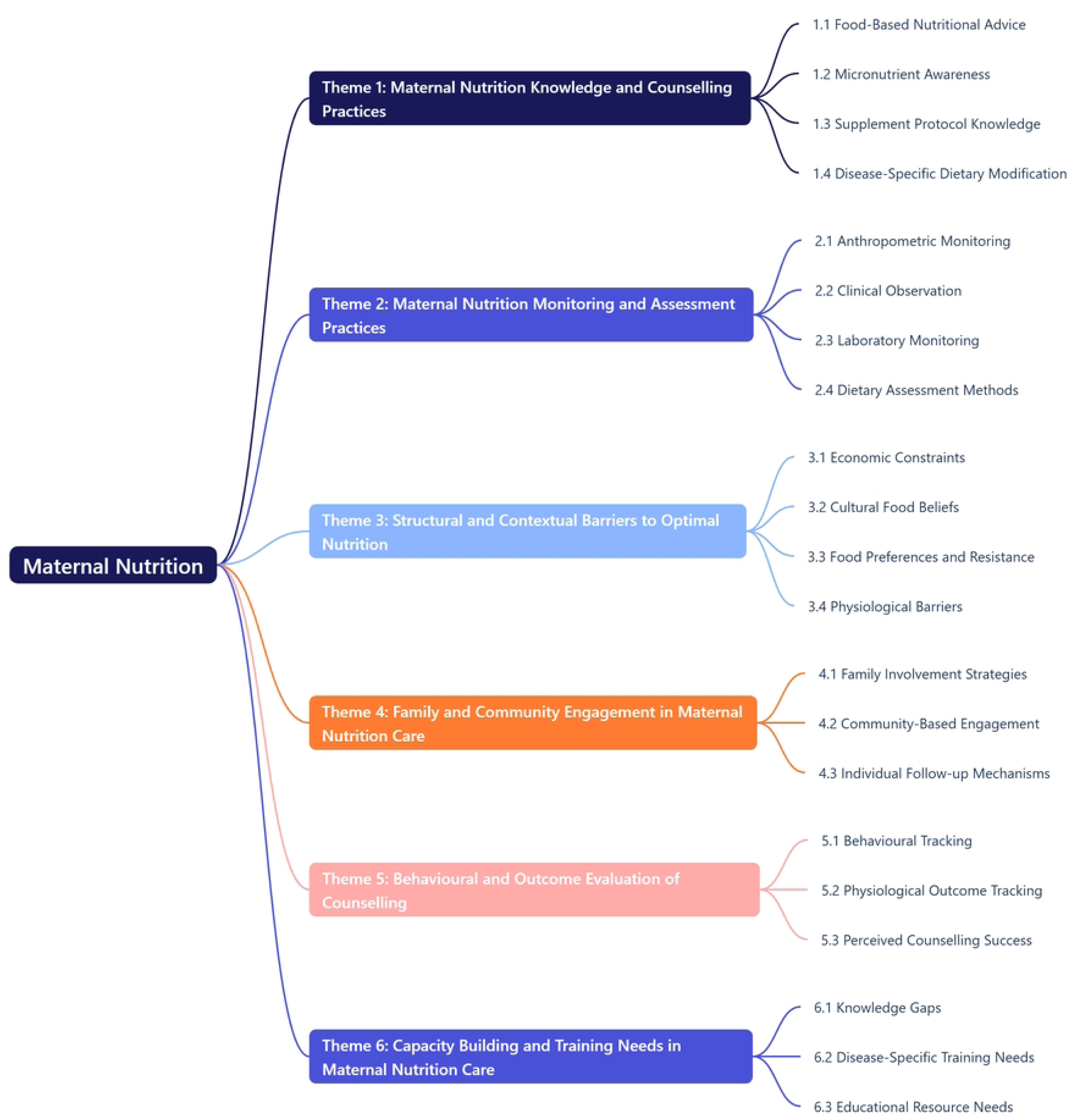
Summary of themes derived from qualitative analysis

**Table 5.**
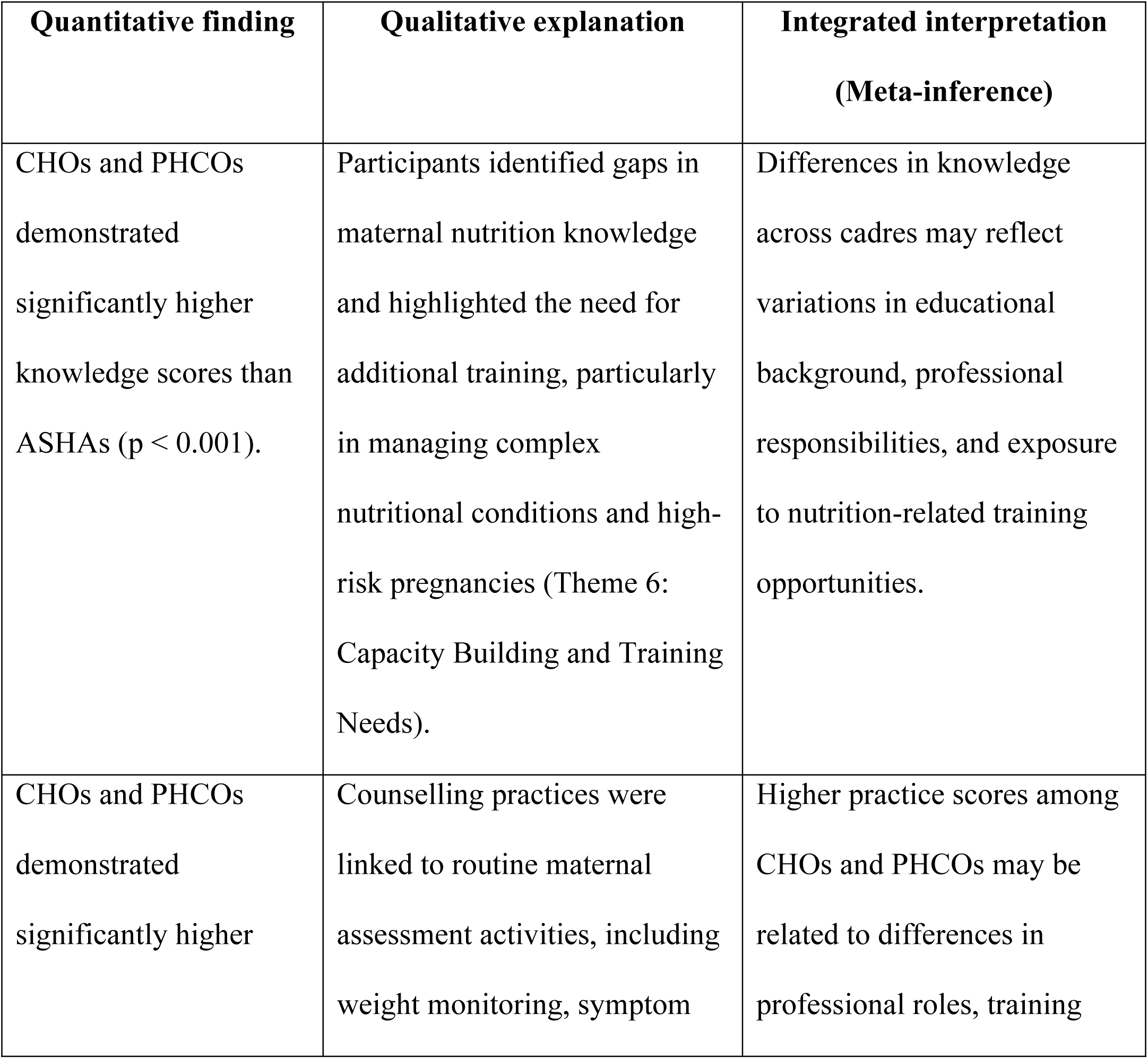
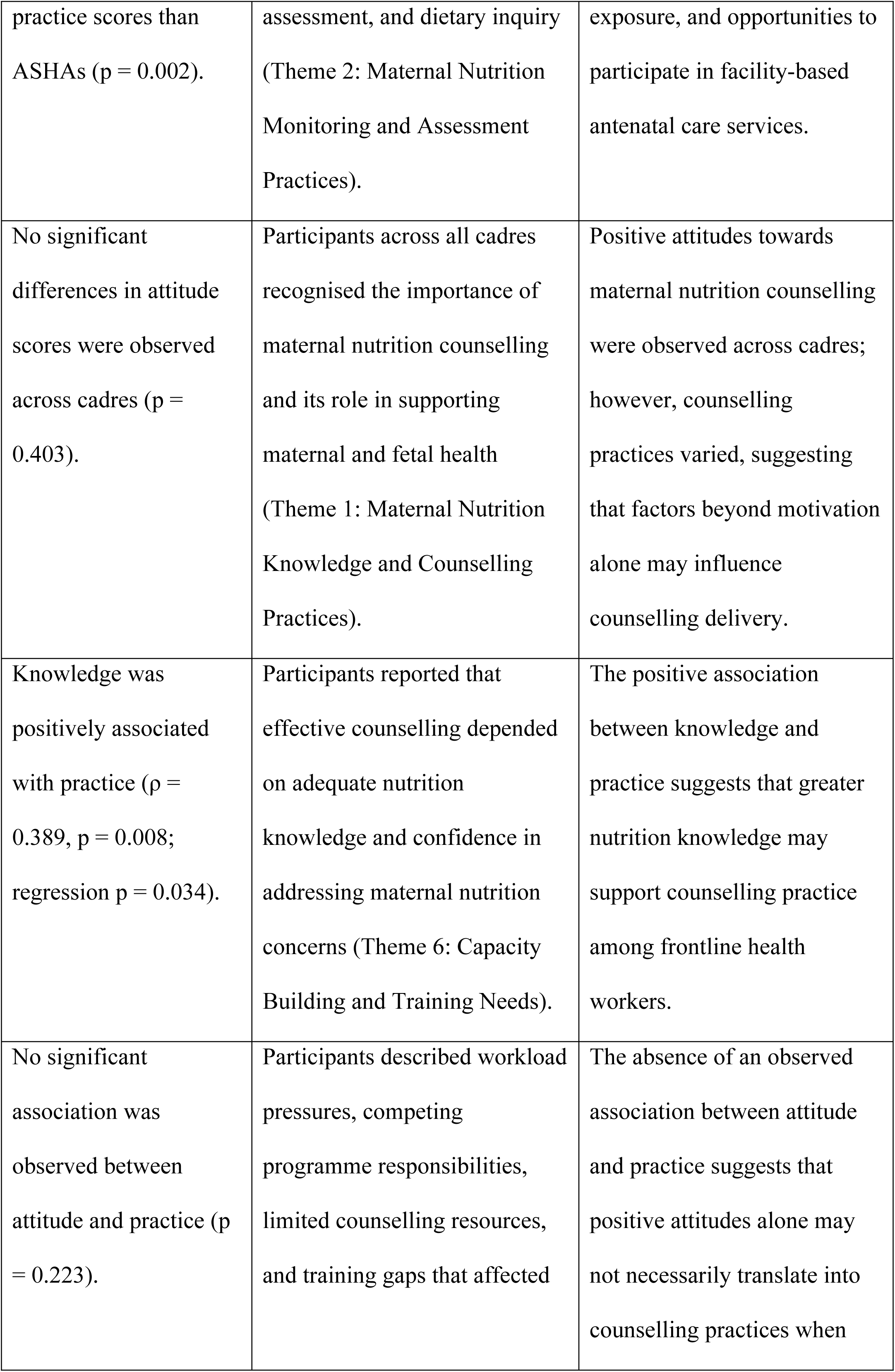
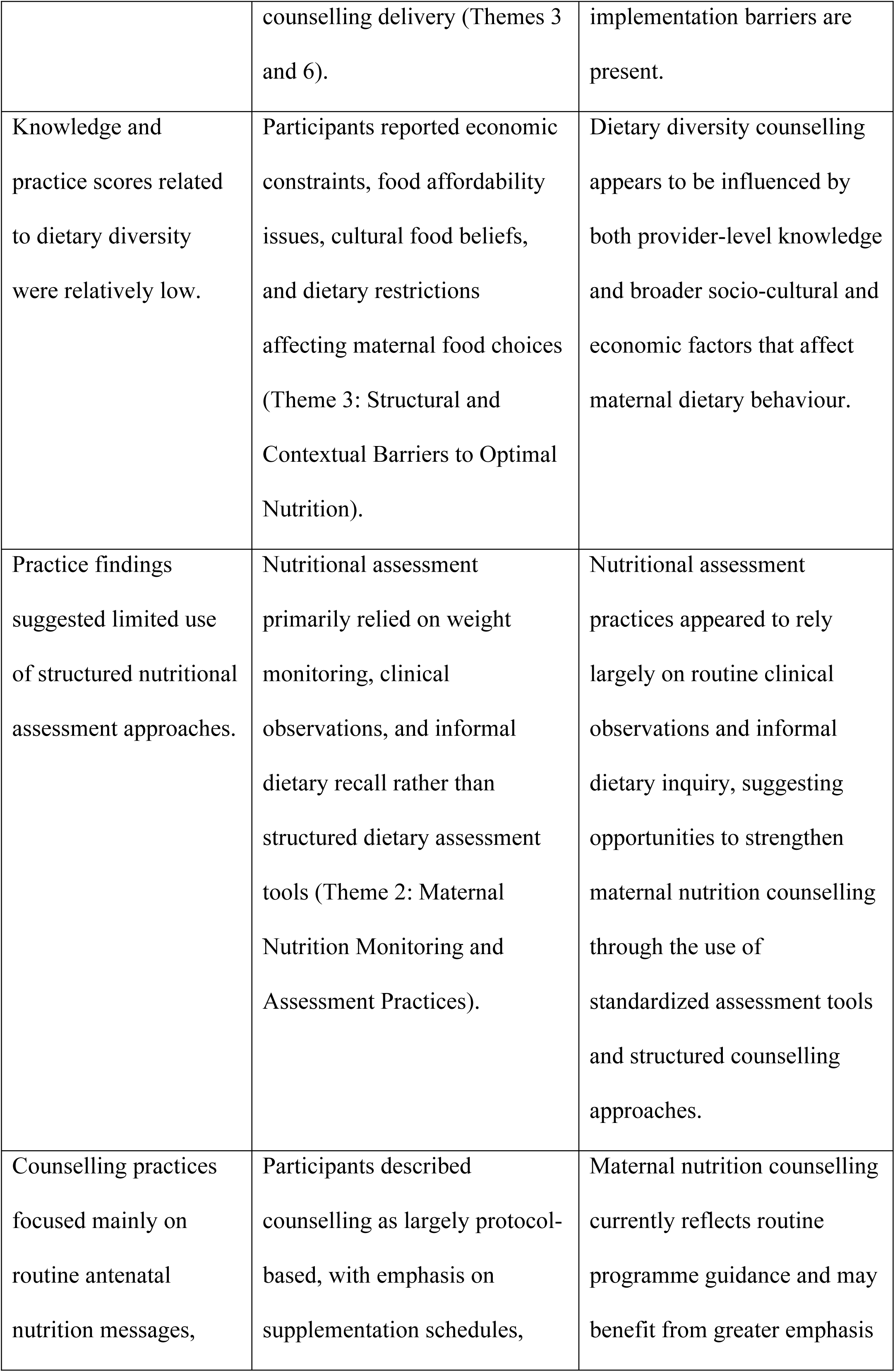
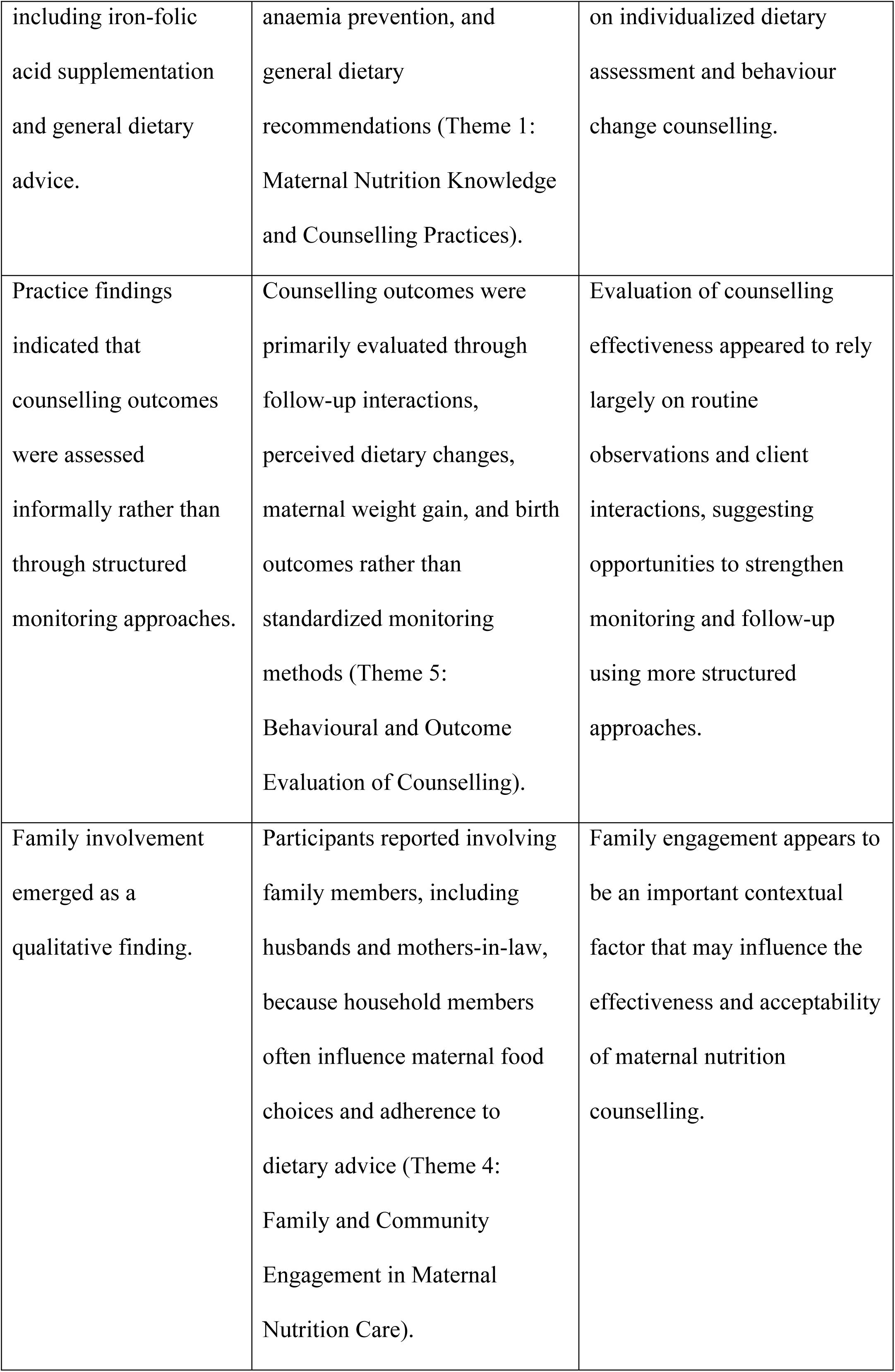
Quantitative and Qualitative findings with meta-inferences.

Table 5 presents a joint display integrating quantitative and qualitative findings to generate meta-inferences regarding maternal nutrition counselling practices among frontline health workers. Quantitative differences in knowledge and practice across cadres were contextualised by qualitative findings related to training exposure, professional roles, and routine service delivery processes. The integrated findings suggest that maternal nutrition counselling is influenced by the interaction of individual competencies, health-system factors, and contextual influences. While positive attitudes towards maternal nutrition counselling were consistently observed across cadres, their translation into practice appeared to be influenced by workload pressures, limited counselling resources, and socio-cultural factors affecting maternal dietary behaviour.

## Discussion

This study used a mixed-methods approach to examine frontline health workers’ knowledge, attitudes, and practices regarding maternal nutrition counselling and to explore contextual factors influencing counselling delivery. The study focused on key maternal nutrition domains, including dietary diversity, micronutrient nutrition, nutritional status assessment, gestational weight gain, and pregnancy-related dietary conditions relevant to contemporary maternal nutrition challenges in India. The findings show gaps in dietary diversity and micronutrient-related knowledge, alongside limited counselling practice, despite uniformly positive attitudes towards maternal nutrition counselling across cadres. This mismatch suggests that, despite being the first point of contact for nutrition care, counselling practices remain primarily designed for earlier nutritional epidemiological contexts rather than the evolving triple burden of malnutrition.

The pattern of limited dietary diversity and micronutrient knowledge observed among FLWs is consistent with previous studies in LMIC primary healthcare settings, where frontline workers often demonstrate general awareness of maternal nutrition but limited depth in applied dietary counselling(24)(26). Prior research from India similarly indicates that antenatal nutrition counselling is often focused on standardised messages, such as iron–folic acid supplementation, rather than comprehensive dietary assessment(27).

The absence of association between attitude and practice aligns with implementation research showing that positive perceptions alone are insufficient to influence behaviour when service delivery constraints are present. In this study, counselling practice remained limited despite uniformly positive attitudes, suggesting that delivery conditions play a more decisive role than motivation alone(28).

The observed differences in knowledge and practice across cadres reflect variation in training exposure and functional roles within the health system. CHOs and PHCOs, who are more closely aligned with facility-based clinical workflows, demonstrated higher knowledge and practice scores compared to ASHAs, who operate primarily at the community level. However, qualitative findings show that counselling practices across all cadres are shaped by similar structural constraints, including limited consultation time, protocol-driven messaging, and lack of structured counselling tools. This suggests that cadre differences primarily reflect variation in system exposure rather than differences in counselling capability.

These findings can be interpreted through an implementation science lens, suggesting that maternal nutrition counselling is shaped not only by individual knowledge and attitudes but also by health system constraints that influence fidelity of guideline delivery within routine antenatal care services. FLWs also reported adapting counselling messages based on affordability, food availability, and cultural food practices. While this reflects contextual responsiveness, it also leads to simplification of dietary diversity guidance, indicating that counselling content is continuously modified at the point of delivery.

Overall, counselling is best understood as a contextually constrained translation of knowledge into practice rather than a linear behavioural outcome. This highlights the need for strengthening not only knowledge acquisition but also practical, system-supported counselling capacity through structured tools and contextualised training approaches. The findings further suggest that strengthening maternal nutrition counselling requires competency-based capacity building that extends beyond factual knowledge acquisition to include counselling skills, dietary assessment, behaviour change communication, and management of emerging pregnancy-related nutritional conditions such as gestational diabetes and excessive gestational weight gain.

## Implications for practice and policy

The findings highlight the need for system-level strengthening of maternal nutrition counselling rather than reliance on training-focused interventions alone. While improving knowledge on dietary diversity and micronutrient needs remains important, the results indicate that knowledge gains alone are insufficient without addressing implementation constraints.

First, introducing structured counselling tools such as visual aids, dietary diversity checklists, and simplified counselling guides may support more consistent delivery of nutrition messages within time-limited consultations. This is consistent with United Nations Children’s Fund(UNICEF)recommendations that effective nutrition counselling should be supported by practical communication tools and behaviour change resources that facilitate client-centred interactions(29).

Second, counselling time and workload distribution within antenatal care services require attention to enable meaningful nutrition counselling beyond brief protocol-based messaging. Third, strengthening supportive supervision and coordination across cadres may help reduce variation in counselling practices and improve consistency in delivery. Fourth, given the influence of household decision-making and cultural food norms, involving family members in counselling processes may enhance dietary adherence and acceptability of recommendations.

These implications are particularly relevant in LMIC contexts such as India, where FLWs serve as the primary interface for maternal nutrition counselling under resource-constrained conditions.

## Limitations

While the study provides valuable insights, certain limitations should be acknowledged. The cross-sectional design limits causal inference between knowledge, attitudes, and practices, and findings should therefore be interpreted as correlational. The relatively small sample size, particularly within individual cadres, may reduce statistical power and limit generalisability beyond the study setting of Udupi taluk, Karnataka, which may differ in health system structure and sociocultural context. Purposive sampling and recruitment during routine meetings may have introduced selection bias.

In addition, self-reported practice measures may be subject to social desirability bias, potentially leading to overestimation of counselling performance. In the qualitative component, data were obtained from a single focus group discussion involving multiple cadres, which may not fully capture the range of perspectives or ensure thematic saturation across all domains. From an analytical perspective, inclusion of multiple predictors in regression models with a limited sample size may introduce risk of overfitting; findings should therefore be considered exploratory.

Despite these limitations, the sequential explanatory mixed-methods design strengthens the study by integrating quantitative patterns with qualitative insights, enhancing contextual interpretation of the findings.

## Conclusion

This mixed-methods study highlights important gaps in the preparedness of frontline health workers to deliver comprehensive maternal nutrition counselling within the context of India’s evolving triple burden of malnutrition. Although maternal nutrition counselling was widely recognised as an important component of antenatal care, limitations in dietary diversity and maternal nutrition knowledge, together with constrained counselling practices, suggest that current counselling approaches may not be optimally positioned to address the key maternal nutrition challenges, including dietary diversity, micronutrient nutrition, nutritional risk assessment, and pregnancy-related dietary concerns. The integration of quantitative and qualitative findings indicates that counselling performance is shaped by the interaction between individual competencies and the broader service delivery environment. Strengthening maternal nutrition counselling therefore requires implementation-focused approaches that extend beyond knowledge enhancement alone and support the delivery of practical, context-responsive counselling within routine antenatal care. Future research should evaluate strategies to strengthen counselling quality and equip frontline health workers to address dietary diversity, micronutrient nutrition, and emerging pregnancy-related nutritional risks.

## Data Availability

The study protocol has been registered and is publicly available on the Open Science Framework (OSF) (https://doi.org/10.17605/OSF.IO/KYAD5). All data supporting the findings of this study are contained within the article and its supplementary materials. Supplementary materials include the KAP questionnaire, focus group discussion guide, and reporting checklists (STROBE and COREQ), along with additional supporting files to ensure transparency and reproducibility.

## Ethics approval and consent to participate

The study was approved by the Institutional Research and Ethical Committees and registered with the Clinical Trials Registry of India (CTRI). Written informed consent was obtained from all participants before enrollment, and the study was conducted in accordance with the principles of the Declaration of Helsinki.

- Institutional Ethics Committee, Manipal Academy of Higher Education, Manipal (Approval No: IEC1: 379/2024; Approved on: 11 September 2024).
- Clinical Trials Registry – India (CTRI) [Registration Number: CTRI/2024/11/076863; Registered on: 18/11/2024].

## Abbreviations

ANC: Antenatal Care
ASHA: Accredited Social Health Activist
BMI: Body Mass Index
CHO: Community Health Officer
COREQ: Consolidated Criteria for Reporting Qualitative Research
CTRI: Clinical Trials Registry of India
CVI: Content Validity Index
FGD: Focus Group Discussion
FLW: Frontline Health Worker
ICDS: Integrated Child Development Services
IEC: Institutional Ethics Committee
IFA: Iron and Folic Acid
KAP: Knowledge, Attitudes, and Practices
LMIC: Low- and Middle-Income Country
MAHE: Manipal Academy of Higher Education
MDD-W: Minimum Dietary Diversity for Women
MMAT: Mixed Methods Appraisal Tool
NHM: National Health Mission
OSF: Open Science Framework
PHCO: Primary Health Care Officer
STROBE: Strengthening the Reporting of Observational Studies in Epidemiology
UNICEF: United Nations Children’s Fund

## Declarations

## Funding

The authors received no specific funding for this work.

## Competing Interests

The authors declare that they have no competing interests.

## Author Contributions

**Conceptualization:** AMS, CRR, SKB, PB

**Methodology:** AMS, CRR, SKB

**Data curation:** AMS, VJ, ERP, TM, SH

**Formal analysis:** AMS, PB, NPK, RB

**Investigation:** AMS, RB, VJ, ERP, TM

**Qualitative analysis:** NPK, RB

**Statistical analysis:** PB

**Interpretation of data:** AMS, CRR, NPK, SKB, RB

**Writing – original draft:** AMS

**Writing – review & editing:** All authors

**Supervision:** SKB, GAM

**Project administration:** SH

## Acknowledgments

The authors sincerely thank all frontline health workers who participated in this study and generously shared their time and experiences. We gratefully acknowledge the Department of Health and Family Welfare, Udupi District, Karnataka, for granting administrative permission and facilitating the conduct of the study across the selected Primary Health Centres and Urban Primary Health Centres. We also acknowledge the support of the health centre staff and field personnel who assisted with participant recruitment and data collection.

The authors express their gratitude to the subject experts who provided guidance during questionnaire development, validation, and methodological refinement.

The use of ChatGPT (OpenAI) and Grammarly for language editing and improvement of grammar, clarity, and readability is acknowledged. These tools were used solely for language refinement and did not contribute to the study design, data collection, analysis, interpretation of findings, or scientific content of the manuscript. The authors take full responsibility for the content and final version of the manuscript.

## Supporting information

**S1 File. KAP Questionnaire**

**S2 File. Focus Group Discussion Guide**

**S3 File. Coding Framework (Codebook)**

**S4 File. STROBE Checklist**

**S5 File. COREQ Checklist**

## Notes

### Competing Interest Statement

The authors have declared no competing interest.

### Clinical Trial

CTRI/2024/11/076863

### Clinical Protocols

https://doi.org/10.17605/OSF.IO/KYAD5

### Funding Statement

The authors received no specific funding for this work. The funders had no role in study design, data collection and analysis, decision to publish, or preparation of the manuscript.

### Author Declarations

The study was reviewed and approved by the Institutional Ethics Committee of Manipal Academy of Higher Education, Manipal, Karnataka, India (Approval No. IEC1:379/2024 approved on 11 September 2024). Administrative approval was obtained from the Department of Health and Family Welfare Services, Udupi District, Karnataka, prior to data collection. Written informed consent was obtained from all participants before enrolment. The study was conducted in accordance with the principles of the Declaration of Helsinki and was registered with the Clinical Trials Registry–India (CTRI/2024/11/076863).

